# Optimal Non-Pharmaceutical Interventions Considering Limited Healthcare System Capacity and Economic Costs in the Republic of Korea

**DOI:** 10.1101/2023.05.24.23290452

**Authors:** Yuna Lim, Youngsuk Ko, Renier Mendoza, Victoria May P. Mendoza, Jongmin Lee, Eunok Jung

## Abstract

Due to the relatively low severity and fatality rates of the omicron variant of COVID-19, strict non-pharmaceutical interventions (NPIs) with high economic costs may not be necessary. We develop a mathematical model of the COVID-19 outbreak in Korea that considers NPIs, variants, medical capacity, and economic costs. Using optimal control theory, we propose an optimal strategy for the omicron period. To suggest a realistic strategy, we consider limited hospital beds for severe cases and incorporate it as a penalty term in the objective functional using a logistic function. This transforms the constrained problem into an unconstrained one. Given that the solution to the optimal control problem is continuous, we propose the adoption of a sub-optimal control as a more practically implementable alternative. Our study demonstrates how to strategically balance the tradeoff between minimizing the economic cost for NPIs and ensuring that the number of severe cases in hospitals is manageable.

## 1 INTRODUCTION

COVID-19, a severe acute respiratory syndrome caused by SARS-CoV-2 has affected public health and the economy worldwide. Healthcare authorities have been responding to COVID-19 and various variants have emerged since the first outbreak. On Nov 24, 2021, the first reported omicron variant in South Africa spread widely around the world, quickly turning into a dominant strain [1]. This new variant has higher transmissibility compared to the delta variant and evades the immunity induced by prior infections or vaccines, but has lower severity and mortality rates [1, 2]. In response to the omicron wave, the Korean health authority focused on severe cases rather than controlling the number of cases [3]. The measures to mitigate COVID-19 pandemic include non-pharmaceutical interventions (NPIs), pharmaceutical intervention, and vaccine intervention. NPIs such as social distancing and lockdowns were implemented worldwide as the first response to the pandemic due to the absence of vaccines and antivirals. Although NPIs are important measures to slow the spread of COVID-19, it leads to high economic costs and burden [4].

Mathematical modeling has been used to determine intervention strategies to reduce the burden of the COVID-19 pandemic. Yuan et al. developed a compartment model reflecting new testing guidelines, social behavior, features of the omicron variant, PCR testing capacity, self-testing, reopening to prepandemic level, and booster dose coverage on the cases and severe cases in Ontario and Toronto, Canada [5]. Ngonghala et al. studied the transmission dynamics of two SARS-CoV-2 variants of concern (delta and omicron) in the United States, in the presence of COVID-19 interventions using a mathematical model [6]. COVID-19 control strategies considering isolation, detection, and treatment based on a 9-dimensional mathematical model with high-risk and low-risk exposures were studied by Li and Guo [7]. Optimal control strategies through cost-effectiveness analysis for each strategy were also presented in their study. Choi and Shim provided information on optimal social distancing and testing strategies for Korea in reducing the disease burden and delaying the peak of the disease using a compartmental model [8]. Silva et al. applied optimal control theory to a deterministic model for COVID-19 to find a balance between reducing transmission rates and returning to normal life at minimal economic cost [9]. Through an age-structured stochastic model for transmission of SARS-CoV-2, Vattiato el al. assessed the potential impact of an omicron wave in New Zealand, which could inform the government of its response [10]. Zong and Luo developed a seven-compartment model with three age groups and it was established that the spread of COVID-19 could be significantly alleviated through proper population control [11]. Oh, Apio, and Park designed a model considering three vaccination levels and predicted that there would be a rapid rise in cases in Korea for a while due to the omicron variant of COVID-19 until herd immunity was achieved [12]. Ko et al. showed an optimal strategy of vaccine allocation to minimize mortality or incidence in Korea using a heterogeneous COVID-19 transmission model. Through this study, the Korean government’s vaccination priority strategy was scientifically supported [13]. Among these, the optimal control problem studies considered minimizing the number of infected people [7, 8, 9]. Considering various controls, the studies proposed optimal strategies to minimize the number of infected people by designing scenarios with a single control or combined controls. In two studies, however, the number of severe cases in hospitals or limited medical resources available for COVID-19 assistance in the country was not considered [7, 8]. In the study considering the number of hospital beds, they obtained the control strategy by formulating the problem as a constrained problem and indirectly reflecting the number of available beds in the upper bound for the number of infections [9].

Our study distinguishes itself from prior research by taking into account the limited number of hospital beds available for severe cases and directly incorporating this information into the objective functional. As a result, we transform the original constrained problem into an unconstrained one, allowing us to identify an intervention strategy that minimizes the number of severe cases in hospitals, rather than simply minimizing the overall number of infected individuals at minimal cost. This approach represents an important contribution to the field of pandemic response planning, as it recognizes the critical role of hospital capacity in mitigating the impact of infectious diseases on public health. We develop a COVID-19 mathematical model taking into account NPIs, vaccination, booster shot, antiviral drugs, and the variants (non-delta, delta, and omicron). Later, optimal control theory is applied to the model to obtain the optimal NPI strategy after the spread of the omicron variant. In particular, we investigate optimal strategy to minimize cost for NPIs and cost for severe cases while preventing the limited healthcare resources from being overwhelmed. We develop an objective functional by including a constraint term reflecting the number of hospital beds for severe cases, which can be realistically secured in Korea. This method provides an optimal strategy to minimize costs without the number of severe cases in hospitals exceeding the level. Though the proposed optimal solution is the optimal strategy, it is considered to be difficult to implement in reality because the value of NPIs changes every day. In consideration of feasibility, we also find a sub-optimal strategy in which each value of NPIs remains constant at least for a few days and which put the same effort as optimal strategy into NPIs.

## 2 MATERIALS AND METHODS

### 2.1 Mathematical model of COVID-19

We present a model that is used to describe the COVID-19 epidemic considering NPIs and imperfect effectiveness of vaccines in Korea. This model will be the framework for the optimal control problem.

The total population is divided into susceptible (*S*), vaccine-related groups (*V, P, W*_*v*_, *B, W*_*b*_), exposed (*E*), infectious (*I*), isolated (*Q*), who moves to mild cases (*M*) or severe cases (*C*), and recovered (*R*). Vaccination refers to administering the primary doses, which include the first and second doses. We assume that the vaccinated compartment (*V*) consists of individuals whose second dose was received within less than two weeks. If more than 2 weeks have passed since the second dose, then an individual moves to the protected (*P*) compartment. The waned after vaccination (*W*_*v*_) compartment consists of individuals with vaccine effectiveness of waned primary doses. The booster-administered (*B*) and waned after booster (*W*_*b*_) compartments consist of those with increased vaccine effectiveness after getting a booster shot and reduced effectiveness of booster shot due to waning, respectively. Note that the vaccinated groups *V, P, W*_*v*_, *B*, and *W*_*b*_ are all susceptible to infection. Figure 1 shows the flow diagram of a leaky vaccine model [14]. That is, if *e*_*P*_ is the effectiveness of vaccines in *P*, then *e*_*P*_ *P* of *P* are protected against infection and (1 − *e*_*P*_) *P* of *P* can be infected.

**Figure 1:**
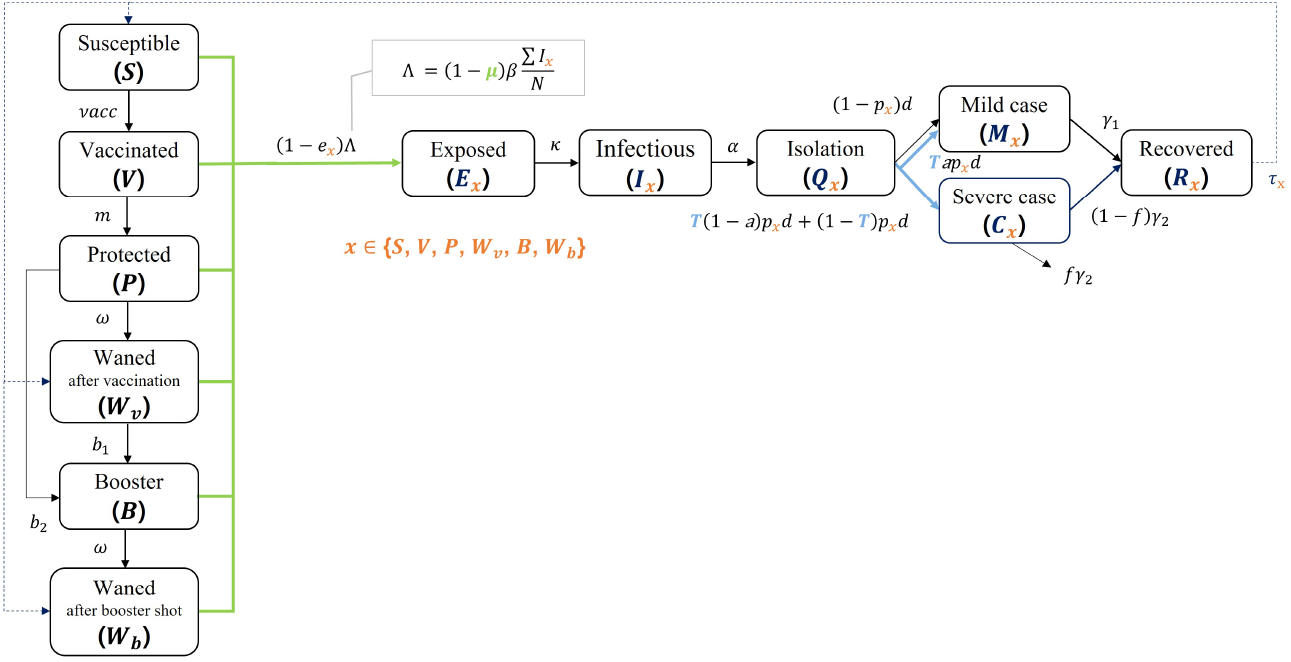
Flow diagram of the mathematical model of COVID-19 epidemic in Korea.

The system of differential equations for the transmission of COVID-19 is as follows:

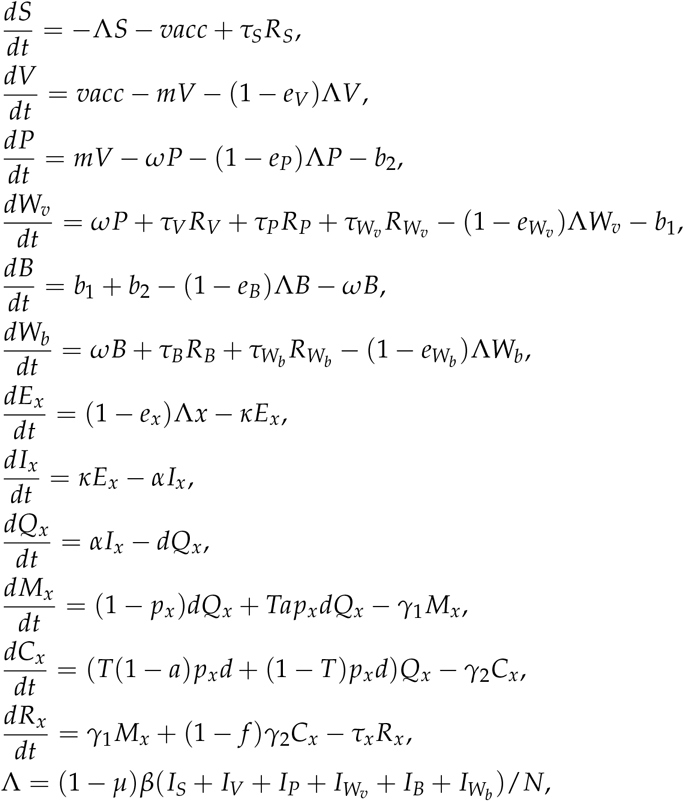

where *N* = *S* + *V* + *P* + *W*_*v*_ + *B* + *W*_*b*_ + *E*_*x*_ + *I*_*x*_ + *R*_*x*_ and *x ∈* {*S, V, P, W*_*v*_, *B, W*_*b*_} .

The factor (1 − *µ*) in Λ represents the reduction in transmission resulting from the NPIs and *β* is the transmission rate. The mean latent period and infectious period are given by 1/*κ* and 1/*α*, respectively. The severity rate *p*_*x*_ depends on the vaccination status of the compartment *x*. The isolation period for mild cases is 1/*γ*_1_ and the duration of hospital stay for severe cases is 1/*γ*_2_. The effectiveness of the primary vaccine or booster shot is expressed as *e*_*x*_. The parameter *T* represents the proportion of the population who are prescribed antiviral drugs. The value of *T* was 0% before 14 Jan 2022 and became 1.45% from 14 Jan to 19 May 2022. Later, it increased to 2.32% from 20 May 2022, as the prescription policy had changed [15, 16]. The effectiveness of antiviral drugs is denoted by *a*. We assume that the drugs are given to compartment *Q*_*x*_ and that the drugs reduce the risk of getting a severe infection by a term *Tap*_*x*_*dQ*_*x*_. Note that *β, κ, α, γ*_1_, *γ*_2_, *p*_*x*_, and *e*_*x*_ are affected by variants. Therefore, we express these parameters as functions of time-dependent detection rates of the delta and the omicron variants. Detailed descriptions of these parameters are given in Appendix A.

The daily number of administered vaccines *vacc* and booster shots *b*_1_ + *b*_2_ were aggregated from the data provided by the Korea Disease Control and Prevention Agency (KDCA). In our model, if the size of *b*_1_ + *b*_2_ is less than the size of *W*_*v*_, then the booster shots are administered only to *W*_*v*_. On the other hand, if the size of *b*_1_ + *b*_2_ is larger than the size of *W*_*v*_, then all individuals in *W*_*v*_ move to *B*, and the difference (*b*_1_ + *b*_2_) − *W*_*v*_ is assumed to be administered to *P*. Thus, there are (*b*_1_ + *b*_2_) − *W*_*v*_ who will move from *P* to *B*. The parameter *m* is the progression rate to immunization after vaccination. The waning rates of vaccine-induced immunity and natural immunity are denoted by *ω* and *τ*_*x*_, respectively. The definitions and values of the parameters are listed in Table 2 of Appendix A.

The timeline in this study is from the start of the community spread of COVID-19 to the post-lifting of social distancing in Korea. We divide this timeline into two periods based on the date when vaccination in Korea started. We set the first period from 16 Feb 2020 to 25 Feb 2021, and the second period from 26 Feb 2021 to 28 Apr 2022. The whole time period spans the predelta-dominant to omicron-dominant periods, which include the time when the dominant delta variant was replaced by omicron. The vaccine-related compartments in the system are excluded in the first period and thus, the flowchart simplifies to a single *SEIQR* path. In accordance with the Korean government’s vaccination strategies, booster shot administration began on 12 Oct 2021. Hence, compartments *B* and *W*_*b*_, and the terms *E*_*x*_, *I*_*x*_, *Q*_*x*_, *M*_*x*_, *C*_*x*_ and *R*_*x*_ with *x* ∈ {*B, W*_*b*_} will only appear from this date.

### 2.2 Least-squares fitting

There are two parameters to be estimated, *µ* and 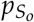, where 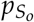 is the severity rate of omicron for the unvaccinated. For estimation of the parameters, we use the least-squares fitting method to minimize the error between the fitted model and the data. From our model, the basic reproductive number of the original strain is expressed as *β*/*α* using the next generation matrix method [17]. The basic reproductive number is estimated at 3.32, i.e., *β*/*α* = 3.32 and *β* = 0.83 [18]. This fixed value of the transmission rate *β* is used for estimating *µ*. We divide the simulation period into three sections: from the beginning of the outbreak to the first vaccine administration (P1: 16 Feb 2020 to 26 Feb 2021), from the first vaccine administration to omicron arrival (P2: 26 Feb 2021 to 30 Nov 2021), and after the first omicron case (P3: 30 Nov 2021 to 28 Apr 2022). In P1, mass gathering-related infections occurred in religious groups, clubs, and nursing homes, and the severe rate was significantly affected by these groups. Hence, severe case data were not used in P1 [19]. On the other hand, confirmed case data in P3 was not used because the COVID-19 test policy was changed and unreported cases cannot be ignored [20]. In Aug 2022, the difference between the proportion of confirmed cases and the rate of testing positive for N antibody was approximately 20%p [21]. Furthermore, we divide each section into different phases according to the NPI-related policies and estimate *µ* on each phase.

### 2.3 Optimal control problem

Our goal is to minimize the cost for NPIs while simultaneously minimizing the number of severe cases. Equivalently, we wish to minimize

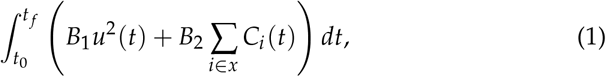

where 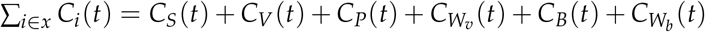 with *x* = {*S, V, P, W*_*v*_, *B, W*_*b*_}. The parameters *t*_0_ and *t* _*f*_ indicate 14 Jan 2022 and 13 Jan 2023, respectively. The term *B*_1_*u*^2^(*t*) is the cost for NPIs and *B*_2_ ∑_*i ∈x*_ *C*_*i*_(*t*) is the cost related to severe cases in hospitals. The cost for NPIs in 2020 was estimated at approximately 30 billion US dollars through a supplementary budget. The weight constant *B*_1_ can be calculated by making *µ* the same form as the cost term *B*_1_*u*^2^(*t*) and setting its value over the period P1 equal to the estimated cost for NPIs in 2020. The two weight constants *B*_1_ and *B*_2_ are set as 221,575 and 1.67, respectively, in consideration of the supplementary budget related to NPIs, the actual cost for severe cases in hospitals, and the relative cost for the two factors.

To ensure that the number of severe cases does not surge at the end of the period, we also added a payoff term, ∑_*i*∈*x*_ *C*_*i*_(*t* _*f*_). We modify (1) and obtain

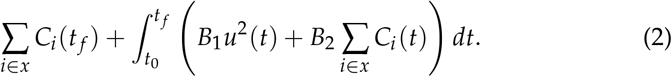

Moreover, we guarantee that the number of severe cases in hospitals does not exceed hospital bed capacity *C*_*max*_. That is,

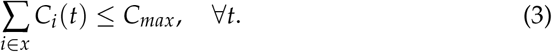

In mid-May 2022, the hospital beds for the treatment of COVID-19 cases were about 80% empty. The government reduced the number of existing beds by half so that patients with other diseases can be accommodated more, reducing the allocation for COVID-19 beds. Reflecting this government’s plan, the value of the *C*_*max*_ is set as 1,500.

We aim to minimize (2) subject to the constraint (3). To do this, we transform this constrained problem to an unconstrained one using a penalty method. We incorporate (3) as a penalty term in (2) and formulate the objective functional *J* given by

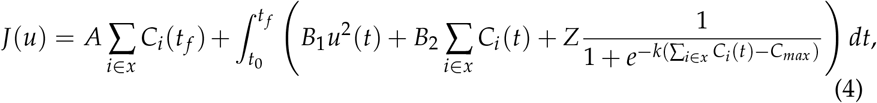

where the constraint is added to the objective functional as a sigmoid function. The sigmoid function is a continuously differentiable approximation of the indicator function, which returns 0 when the number of severe cases in hospitals ∑_*i∈x*_ *C*_*i*_(*t*) does not reach *C*_*max*_, and 1 whenever ∑_*i ∈x*_ *C*_*i*_(*t*) exceeds *C*_*max*_. The weight constant *Z* of the constraint term is set to a very large value (1,000,000). The coefficient *k*, associated with the slope at *C*_*max*_, is set to 0.1. We seek the optimal control *u*^∗^, which minimizes the objective functional, satisfying

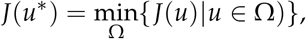

where 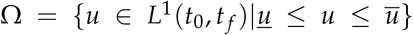. The intensity of NPIs in 2022 is not expected to be higher than in 2021 because Korea implemented a gradual recovery policy, which aims to ease the restrictions. So, we set the upper bound ū as the maximum value of the *µ* values estimated in each period. When all NPIs are lifted, then *µ* = 0, and so, we set the lower bound 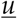 to 0.

We solve the optimal control problem starting from 14 Jan 2022, the date when antiviral drugs were first introduced in Korea, using the forward-backward sweep method [22]. We consider a time-dependent control *u*(*t*) that represents the intensity of efforts to reduce the spread of infection by implementing NPIs. In the model, *u*(*t*) replaces *µ* starting on 14 Jan 2022. The characterization of the optimal control is in Appendix B. Furthermore, the minimizer of (4) is compared to the optimal control without constraint and the *µ* estimated in 2022. Since social distancing has been lifted since 18 Apr 2022, it is assumed that the *µ* value after 28 Apr 2022 is the same as the *µ* value estimated from 18 Apr to 28 Apr 2022.

Because the solution *u*^∗^ of the optimal control problem is a continuous function, the corresponding strategy is time-dependent and gives a value that differs every moment. This is unrealistic and will be impossible to implement. To address this, an approach is to find an alternative control, which is computed by formulating a piecewise control from the obtained optimal control [23]. The detailed description of how this sub-optimal piecewise control is formulated is discussed in detail in Appendix C.

## 3 RESULT

### 3.1 Parameter estimation

Figure 2 shows the model (black solid curves) fitted to the cumulative confirmed cases data (blue circles) and cumulative severe cases data (red circles). The model was fitted to the confirmed cases data from 16 Feb 2020 to 26 Feb 2021, and to severe cases data from 30 Nov 2021 to 28 Apr 2022. From 26 Feb to 30 Nov 2021, both the confirmed cases and severe cases data were used in the model fitting. The maximum, minimum, and average values of the fitted parameter *µ* are 0.71 (from 26 Feb 2021 to 27 Apr 2021), 0.117 (from 18 Apr 2022 to 28 Apr 2022), and 0.459, respectively. The parameter estimates are listed in Table 3 in Appendix A. The estimated value of 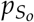 was 0.00053, which is 2.36% of the severity rate of the delta variant. The parameter *B*_1_, which is the weight constant for the NPIs, is calculated as 167,947,693 US dollars using the estimated values of *µ*.

**Figure 2:**
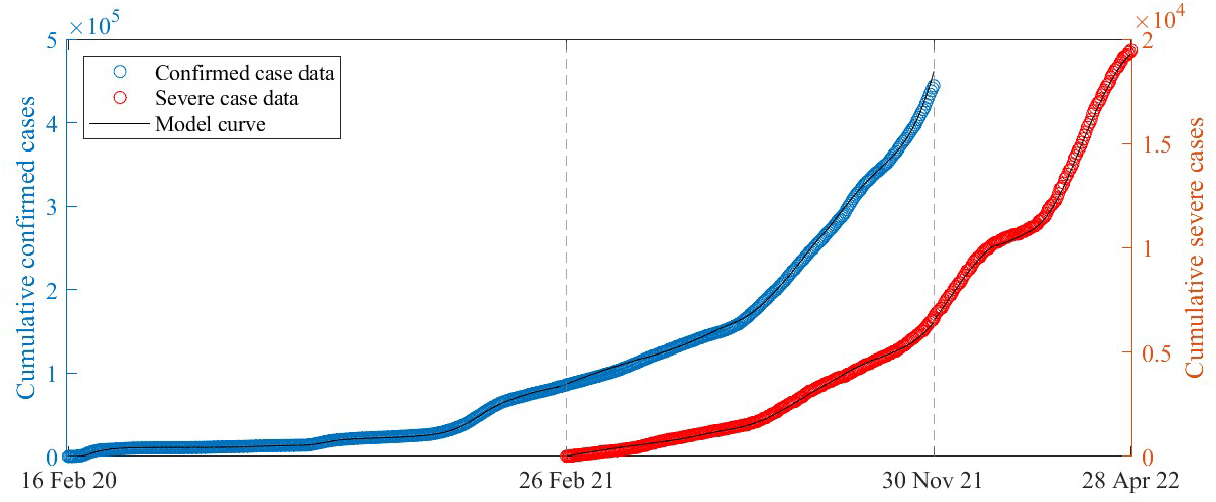
The best model fit (black curves) to the data from 16 Feb 2020 to 28 Apr 2022. Both the confirmed cases (blue circles) and severe cases (red circles) data were used in the fitting from 26 Feb 2021 to 30 Nov 2021, while only the confirmed (severe) cases data were used from 16 Feb 2020 to 26 Feb 2021 (30 Nov 2021 to 28 Apr 2022).

### 3.2 Characteristics of optimal control

#### Optimal controls with and without constraint and NPIs control estimated

Figure 3 displays the optimal and estimated values of the NPIs, the cumulative confirmed cases, and severe cases in hospitals. The blue and red solid curves indicate the results of the optimal controls with and without constraint on the hospital bed capacity, respectively. The black dashed curve represents the results using the estimated values of *µ*. If the constraint is included in the objective functional (blue solid line), there is a period when the NPIs control is enforced relatively strongly. The optimal control rises to 0.61 (39th day to 44th day) and then drops to almost zero afterwards. However, if there is no constraint (red solid line), the value of the optimal NPIs control is close to zero on the entire period. Consequently, since the NPIs control is minimally implemented, a relatively large number of confirmed cases occur, and the number of severe cases in hospitals exceeds *C*_*max*_. The estimated *µ* values, which were fitted to follow the data, vary between 0.117 and 0.61. The number of cumulative confirmed cases corresponding to the estimated *µ* is relatively lower than the other two cases, and the number of severe cases in hospitals is less than *C*_*max*_. The optimal control resulting from the constrained objective functional is relatively lower than the estimated *µ* except from the 26th to 50th days. As a result, the number of cumulative confirmed cases corresponding to the optimal control with constraint is relatively larger, but the number of severe cases in hospitals is still below *C*_*max*_.

**Figure 3:**
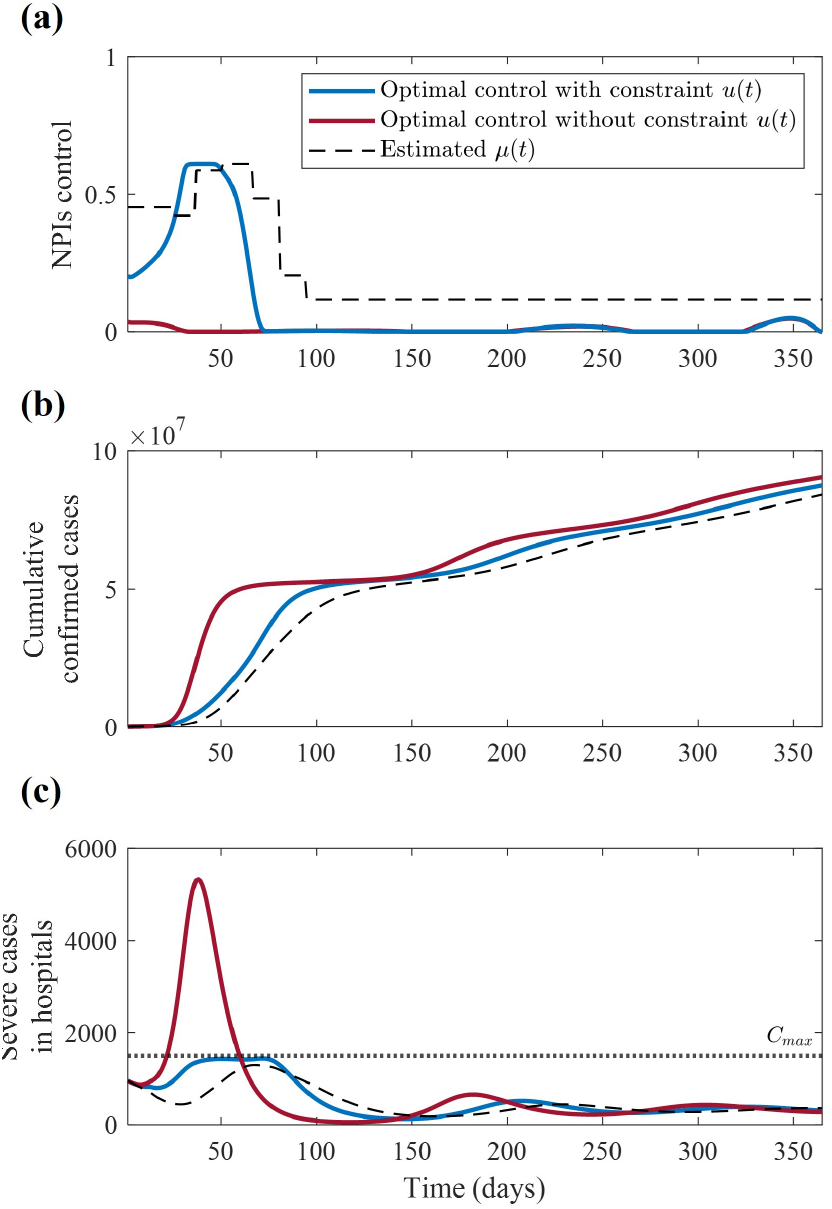
The optimal controls when the constraint is included (blue solid curve) and excluded (red solid curve) in the objective functional, and the estimated values of the NPIs implemented in 2022 (black dashed curve). From the top, (a) NPIs control, (b) corresponding cumulative confirmed cases, and (c) corresponding severe cases in hospitals are shown. In panel (c), the black dotted line indicates the manageable level *C*_*max*_ of severe cases in hospitals in Korea.

#### Alternative controls

Figure 4 shows the (a) optimal, average, and sub-optimal controls, (b) cumulative confirmed cases, and (c) severe cases in hospitals. In panel (a) of Figure 4, the gray dotted line indicates the detection rate of the omicron variant. Based on the values of the optimal control, the period is divided into 5 intervals. The average control is calculated as the average value of the optimal control on each interval. However, when the average control is applied, the number of severe cases in hospitals in the first interval exceeds *C*_*max*_. Therefore, the first interval is divided further into 3 sub-intervals based on the intersections of the plots of the optimal and average controls in the first interval. The details for obtaining the sub-optimal control are provided in Appendix C of the supplementary material. The areas under the curves of the average and sub-optimal controls are equal to the area under the curve of the optimal control. The numbers of confirmed and severe cases in hospitals are similar when the sub-optimal and optimal controls are applied. Since the 71st day, all NPIs controls are close to zero. The number of severe cases in hospitals corresponding to all NPIs controls is within *C*_*max*_ after the first peak.

**Figure 4:**
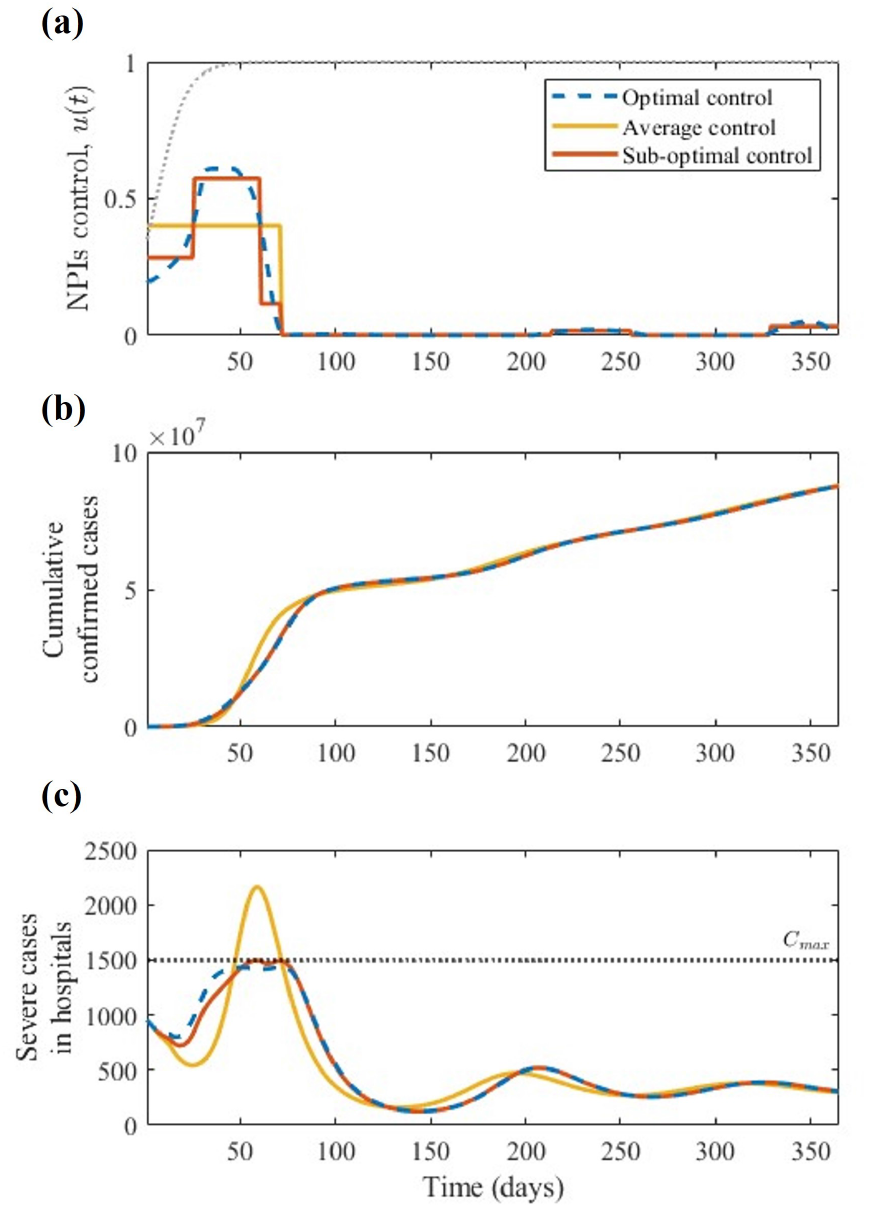
The optimal (blue solid curve), average (yellow solid curve), and suboptimal controls (orange solid curve). Panel (a) shows the three NPIs controls. The gray dotted curve indicates the detection rate of the omicron variant. Panels (b) and (c) show the corresponding cumulative confirmed and severe cases, respectively. The black dotted line in panel (c) shows the manageable level *C*_*max*_ of severe cases in hospitals in Korea.

## 4 DISCUSSION

The values of *µ* can be interpreted as quantified effects of NPIs against disease spread, except in some periods related to mass gathering events [24, 25, 26]. For example, social distancing level 4, which prohibits gatherings of more than three people after 6 p.m., events and gatherings other than one-person protest, and face-to-face religious activities, could reduce the force of infection by 65%. By estimating the *µ* values during the entire period, an upper bound for the control is determined. The estimated value of *µ* until the start of vaccination was used to tune the cost-related parameter *B*_1_, which appeared in the optimal control problem. This approach can be easily modified and applied to other countries or diseases.

Intensified NPIs can cause significant economic burden [27]. We also found that the estimated weight of NPIs (*B*_1_) is 132,680 times more than the cost for a severe case (*B*_2_). Hence, in the absence of the constraint, NPIs control is less favored as a way to reduce costs. No NPIs may lead to more incidence and severe cases in hospitals, and puts a strain in the healthcare system. The red solid line in Figure 3 shows that the optimal control for the problem without constraint could not be realistically implemented, considering the healthcare system capacity. The number of severe cases in hospitals was suppressed below *C*_*max*_ by including a constraint in objective functional. We also estimated the reduction in transmission due to NPIs (*µ*) implemented from 14 Jan to 28 Apr 2022. The trend of the estimated *µ* value in each period in Figure 3 is similar to the change of social distancing policies (in Table 3).

Comparing the number of cumulative confirmed cases and severe cases in hospitals corresponding to the optimal and estimated *µ* in Figure 3, it can be seen that Korea strongly implemented NPIs in the first half of 2022. Although there are relatively more incidence cases if the optimal control is applied, the number of severe cases in hospitals could be kept below *C*_*max*_ with more eased NPIs.

Subsequently, we found a less costly sub-optimal control, obtained from the optimal control. In Figure 4 (a), the area below each curve is the same, which means that the effort involved in NPIs is the same. Even though the area below the curves of the average and sub-optimal controls from the 1st day to 71st day are the same, there are differences in the sizes of the severe cases in hospitals corresponding to each control during this period. This means that a strategic implementation of NPIs is necessary from the 1st day to 71st day, the period when the detection rate of the omicron variant increases to 100%. The obtained time-dependent optimal control shows the importance of prompt application of the enhanced NPIs when omicron rapidly became the dominant variant.

Limitations of this research are as follows. First, we did not consider unreported cases, which have been suspected to exist after the omicron wave because the proportion of the population who have infection-induced antibody exceeded the proportion of those who were confirmed to have COVID19 [21]. Therefore, the number of cumulative confirmed cases simulated from our model might differ from the actual data. Second, we did not consider a long-lasting partial immunity induced only by infection but assumed that the unvaccinated host whose immunity from infection had waned has the same immunity as those who never got infected. Re-infection considering partial immunity and unreported cases will be included in the future work.

## 5 CONCLUSION

In this study, we obtained an optimal control which was designed to minimize costs while suppressing the number of severe cases in hospitals below the manageable level in Korea. To do this, we employed an innovative methodology that transforms the constrained problem into an unconstrained one, using a logistic function and incorporating the limited hospital beds as a penalty term in the objective functional. Model simulations using the optimal control emphasize that the number of severe cases in hospitals could be reduced to the manageable level with less effort and cost for NPIs than in practice.

Based on the optimal control, we also suggested the more practical suboptimal control. We found that the number of severe cases in hospitals could be under the manageable level with less effort and cost. Furthermore, NPIs should be intensified during the period when the omicron variant becomes dominant. The framework of this study can be modified to consider other variants, or even another emerging infectious disease which can appear in the future, and suggest appropriate level of NPIs to secure the healthcare system.

## Data Availability

All data produced in the present study are available upon reasonable request to the authors

## 6 Acknowledgements

This paper is supported by the Korea National Research Foundation (NRF) grant funded by the Korean government (MEST) (NRF-2021M3E5E308120711). This paper is also supported by the Korea National Research Foundation (NRF) grant funded by the Korean government (MEST) (NRF-2021R1A2C100448711).

## Appendices

### A Model parameter setting

The proportions of the variants are modeled using a logistic function that follows the reported detection rates of the delta and omicron variants among the infections [28, 29, 30]. The proportions of each variant over time are formulated

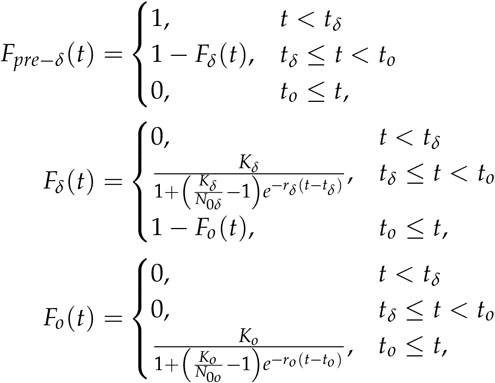

where *N*_0*δ*_ (*N*_0*o*_) is the initial detection rate of delta (omicron) which was aggregated from the data [31, 32], *K*_*δ*_ (*K*_*o*_) is the upper limit of the proportion of delta (omicron) which we set to 1, *r*_*δ*_ (*r*_*o*_) is the growth rate of the proportion of delta (omicron) among the infections, and *t*_*δ*_ (*t*_*o*_) is the initial detection time of the delta (omicron) variant. The parameters *r*_*δ*_ and *r*_*o*_ are computed using the values of *N*_0*δ*_, *K*_*δ*_, *t*_*δ*_, *N*_0*o*_, *K*_*o*_, and *t*_*o*_. Their values are 0.091 and 0.127, respectively.

If *X* is a parameter which is affected by the variants, then its value is formulated as a function of the detection rate of variants (*F*_*pre − δ*_(*t*), *F*_*δ*_(*t*), *F*_*o*_(*t*)) and variant-specific parameters (*X*_*pre− δ*_,*X*_*δ*_,*X*_*o*_). The parameter *X* is expressed as follows,

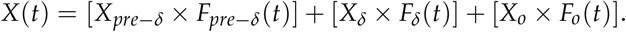

In our study, the effectiveness of booster shots against the pre-delta variant was not considered because the booster administration started only during the delta-dominant period [33, 34]. Because of a new isolation policy from 26 Jan 2022 [35], *γ*_1_ and *γ*_2_ changed to 1/7 and 1/10 from this date, respectively. The variant-dependent parameters are listed in Table 1 and the rest of the parameters are listed in Table 2. The phase-dependent *µ* and the estimates are listed in Table 3. We see in Table 3 the values of *µ* on several periods, divided according to the social distancing policy. In our model, *β* represents the transmission rate in the community and not specific to groups. A significantly higher reproductive number than the basic reproductive number were observed in two periods in 2020, where the *µ* values were estimated to be negative [36, 18]. For example, in the period from 16 Feb 2020 to 28 Feb 2020, the estimated value of *µ* is −0.939, which translates to an effective reproductive number, expressed as (1 − *µ*) 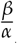, of 6.437. Note that these negative *µ* values are ignored during the estimation *B*_1_.

**Table 1:** Parameters and definitions used in the model according to variants

| Symbol | Description | Pre-delta | Delta | Omicron | Reference |
| --- | --- | --- | --- | --- | --- |
| $\kappa$ | Progression rate | 1/4 | 1/2 | 1/2 | [37, 38, 39] |
| $\alpha$ | Isolation rate | 1/4 | 1/6 | 1/6 | [40, 41] |
| $\beta$ | Transmission rate without interventions | 0.83 | 0.85 | 1.583 | [18, 42, 43] |
| $\gamma_1$ | Recovery rate of mild case | 1/14.7 | 1/11.9 | 1/10 | [44] |
| $\gamma_2$ | Recovery rate of severe case | 1/17.5 | 1/14.3 | 1/12.6 | [44] |
| $e_v$ | Effectiveness of first shot | 0.625 | 0.47 | 0.37 | [45, 46, 47] |
| $e_p$ | Effectiveness of second shot | 0.833 | 0.893 | 0.626 | [45, 46, 47] |
| $e_{W_v}$ | Waned effectiveness of primary vaccination | 0.662 | 0.605 | 0.074 | [45, 46, 47] |
| $e_b$ | Effectiveness of booster shot | - | 0.858 | 0.72 | [45, 46, 47] |
| $e_{W_b}$ | Waned effectiveness of booster shot | - | 0.751 | 0.331 | [45, 46, 47] |
| $p_S$ | Severe rate of the unvaccinated | 0.0183 | 0.022 | 0.0005 | [48] |
| $p_P$ | Severe rate of people with effectiveness of primary vaccine | 0.0031 | 0.0037 | 0.0002 | [48, 49] |
| $p_{W_v}$ | Severe rate of people with waned effectiveness of primary vaccine | 0.0042 | 0.0051 | 0.0002 | [48, 49] |
| $p_B$ | Severe rate of people with effectiveness of booster shot | 0.0011 | 0.0013 | 0.0001 | [48, 49] |
| $p_{W_b}$ | Severe rate of people with waned effectiveness of booster shot | - | 0.0025 | 0.0001 | [48, 49] |

**Table 2:** Parameters and definitions used in the model

| Symbol | Description | Value | Reference |
| --- | --- | --- | --- |
| $1/d$ | Severity classification period | 0.5 | Assumed |
| $f$ | Case fatality rate | 0.0078 | [50] |
| $\tau_S$ | Waning rate of natural immunity of the unvaccinated | 1/120 | [51, 52, 53] |
| $\tau_x$ ($x \in \{V, P, W_v, B, W_b\}$) | Waning rate of natural immunity of the vaccinated | 1/480 | [54] |
| $m$ | Progression rate from compartment V to compartment P | 1/48.51 | [55, 56] |
| $\omega$ | Waning rate of vaccine-induced immunity | 1/180 | [57, 58] |

### B Characterization of optimal control

An optimal control and the corresponding states should satisfy the necessary conditions derived from Pontryagin’s maximum principle [59]. The Hamiltonian of our model is defined as follows,

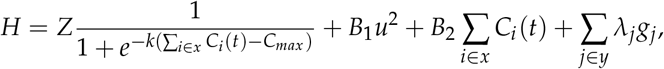

where *λ*_*j*_ is the adjoint variable and *g*_*j*_ is the right hand side of each differential equation for the state variables. Note that *x* and *y* indicate the set of state variables related to the vaccination status and the set of all state variables used in the model, respectively, i.e., *x* = {*S, V, P, W*_*v*_, *B, W*_*b*_} and 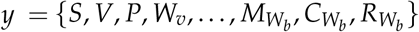. The necessary conditions are obtained from the Hamiltonian *H* as follows [22]:

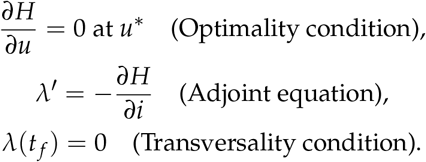

We obtain the adjoint equations by differentiating the Hamiltonian *H* with respect to each state variable *i* ∈ *x*:

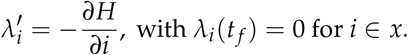

**Table 3:** *µ* value by period

| Value | Period | Value | Period |
| --- | --- | --- | --- |
| -0.939 | 16 Feb 2020 to 28 Feb 2020 | 0.640 | 28 Apr 2021 ~ 30 Jun 2021 |
| 0.871 | 29 Feb 2020 to 21 Mar 2020 | 0.490 | 1 Jul 2021 ~ 11 Jul 2021 |
| 0.878 | 22 Mar 2020 to 19 Apr 2020 | 0.580 | 12 Jul 2021 ~ 26 Jul 2021 |
| 0.881 | 20 Apr 2020 to 8 May 2020 | 0.650 | 27 Jul 2021 ~ 7 Sep 2021 |
| 0.483 | 9 May 2020 to 23 Jun 2020 | 0.530 | 6 Sep 2021 ~ 28 Sep 2021 |
| 0.723 | 24 Jun 2020 to 8 Aug 2020 | 0.513 | 29 Sep 2021 ~ 17 Oct 2021 |
| -0.212 | 9 Aug 2020 to 15 Aug 2020 | 0.378 | 18 Oct 2021 ~ 31 Oct 2021 |
| 0.616 | 16 Aug 2020 to 29 Aug 2020 | 0.191 | 1 Nov 2021 ~ 30 Nov 2021 |
| 0.879 | 30 Aug 2020 to 13 Sep 2020 | 0.378 | 1 Dec 2021 ~ 14 Dec 2021 |
| 0.747 | 14 Sep 2020 to 27 Sep 2020 | 0.380 | 15 Dec 2021 ~ 5 Jan 2022 |
| 0.747 | 28 Sep 2020 to 11 Oct 2020 | 0.400 | 6 Jan 2022 ~ 13 Jan 2022 |
| 0.590 | 12 Oct 2020 to 18 Nov 2020 | 0.453 | 14 Jan 2022 ~ 6 Feb 2022 |
| 0.480 | 19 Nov 2020 to 30 Nov 2020 | 0.422 | 7 Feb 2022 ~ 18 Feb 2022 |
| 0.632 | 1 Dec 2020 to 22 Dec 2020 | 0.586 | 19 Feb 2022 ~ 4 Mar 2022 |
| 0.776 | 23 Dec 2020 to 3 Jan 2021 | 0.610 | 5 Mar 2022 ~ 20 Mar 2022 |
| 0.747 | 4 Jan 2021 to 14 Feb 2021 | 0.485 | 21 Mar 2022 ~ 17 Apr 2022 |
| 0.529 | 15 Feb 2021 to 25 Feb 2021 | 0.204 | 4 Apr 2022 ~ 17 Apr 2022 |
| 0.710 | 26 Feb 2021 to 27 Apr 2021 | 0.117 | 18 Apr 2022 ~ 28 Apr 2022 |

The optimal control *u*^∗^ is derived using the optimality condition at *u*^∗^ on the set Ω = {*u* ∈ *L*^1^(*t*_0_, *t* _*f*_)|0 ≤ *u* ≤ 0.61}.

### C Computation of sub-optimal control

Sub-optimal control should satisfy the following conditions: (1) optimal control *u*^∗^ and sub-optimal control have the same average value, (2) the severe bed capacity is not exceeded, and (3) the cost is the lowest among all possible piecewise combinations.

We formulated the piecewise control by first dividing the period into different intervals, which are chosen whenever the optimal values are less than 10^−2^ or not. Then the average values of *u*^∗^ are calculated for each interval. This is how the average control is obtained. However, this does not guaranteed condition (2). So in our problem, the first interval is divided into 3 sub-intervals based on the intersections of the optimal control *u*^∗^ and average control. That is,

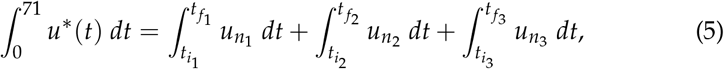

where 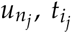, and 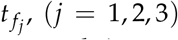, indicate NPIs value, the initial date, and the end date in the *j*th sub-interval, respectively. We discretize 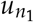 and 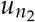 with step size set to 0.01 and found a total of 17966 possible combinations. The expression in (5) is utilized to determine 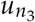 for each 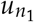 and 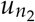. Using the second condition, the number of combinations is reduced to only 724 combinations. Finally, the last condition is used to determine the sub-optimal control by finding the combination that will yield the least value for objective functional *J*. Panels (a) and (b) of Figure 5 illustrate the total cost and the maximum number of severe cases, respectively, for varying values of 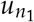 and 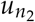. The red contour indicates the combinations of 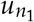 and 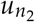 such that the maximum value of the number of severe cases is exactly 1,500. This means that the interior of the red curve represents all the combinations of 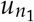 and 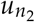 that will yield to severe cases not exceeding 1,500. The white star represents the lowest total cost.

**Figure 5:**
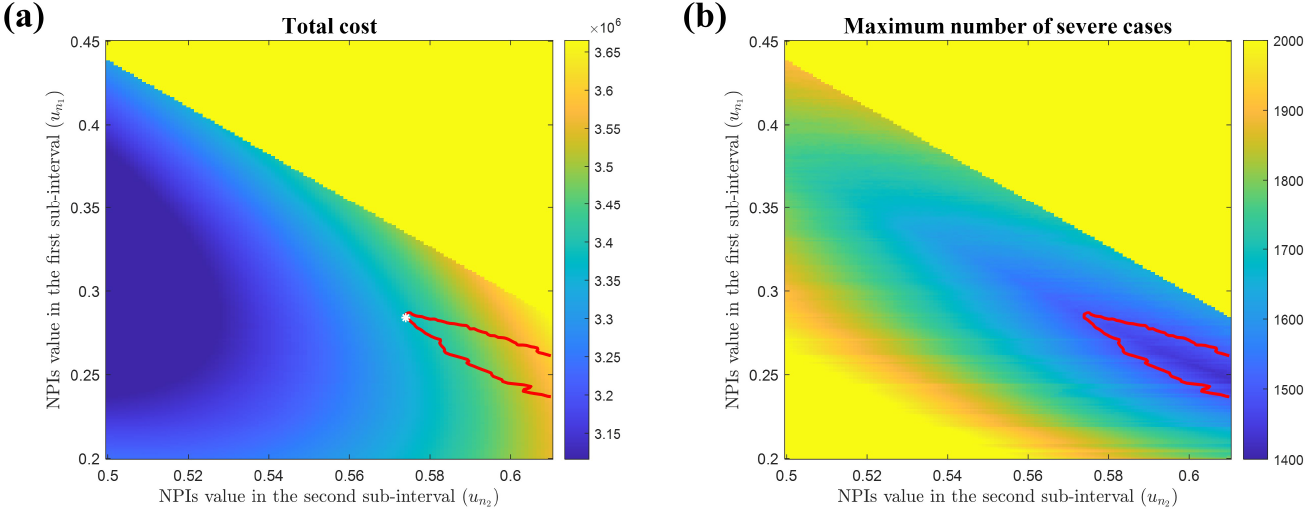
The heat map of the total cost and the maximum number of severe cases for varying values of 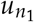 and 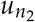 . The red contour indicates the maximum value of the number of severe cases in hospitals, which is 1,500, and the white star represents the sub-control with the least objective functional value.

